# Progress on International Health Regulations core capacities in the Western Pacific Region

**DOI:** 10.1101/2025.03.02.25322262

**Authors:** Kai Xiao, Qiu Yi Khut, Phuong Nam Nguyen, Ariuntuya Ochirpurev, Sean Casey, Jessica Kayamori Lopes, Gina Samaan

## Abstract

The International Health Regulations (IHR 2005) are binding upon Member States of the World Health Organization (WHO), requiring them to build and maintain capacities across critical domains to prevent, detect and respond to public health threats. In an analysis of 15 IHR core capacity scores reported by States Parties in the Western Pacific Region from 2021–2023, the average regional scores increased from 68% in 2021 to 72% in 2022, then declined to 66% in 2023. Seven States Parties maintained consistently strong scores (≥85%), whereas nine exhibited fluctuations of at least 10 percentage points. Categorization of States Parties into three groups based on geographic and economic characteristics highlighted that core capacities such as financing, food safety and zoonotic disease control were areas requiring additional support, particularly among Pacific island States Parties. Low and middle income States Parties also reported notable gaps in financing and infection prevention and control. These findings underscore the importance of strategically establishing or designating a National IHR Authority (NIA), which is a key amendment to the IHR implemented in 2024. Beyond technical improvements, a strong NIA can drive multisectoral collaboration, help mobilize resources and streamline decision making. Additionally, establishing a regional forum for health emergencies could enhance political commitment and promote joint actions, strengthening collective resilience. In turn, this fosters more resilient preparedness and response measures that address the diversity epidemiological, economic and geographical contexts in the region, thereby strengthening overall IHR implementation.

## INTRODUCTION

The International Health Regulations (IHR 2005) constitute a legally binding international instrument for States Parties, which include all Member States of the World Health Organization (WHO).^1^ States Parties are obligated to establish, strengthen and maintain the necessary core health capacities across sectors for rapid detection, timely reporting and effective response to public health risks and emergencies, thereby contributing to global health security.

Since 2005, States Parties have made significant strides in enhancing IHR core capacities. However, the COVID-19 pandemic revealed vulnerabilities in global health systems, including preparedness gaps, delayed reporting and insufficient coordination across relevant sectors and borders. These issues underscore the need to further strengthen core capacities and establish more robust mechanisms for multisectoral coordination to secure their full implementation. In response, Member States commenced a process to amend the IHR (2005) to address these deficiencies.

Since 2022, the Member State-led Working Group on Amendments to the International Health Regulations has reviewed over 300 proposed changes to the IHR in light of learnings from COVID-19. A set of amendments was adopted after 2 years of negotiation by the Seventy-seventh World Health Assembly in June 2024. The amendments focus on enhanced coordination, capacity building and rapid response mechanisms across all levels of the health security architecture.^2^ By fulfilling these new obligations, States Parties will play a vital role in strengthening global health security and preventing the international spread of diseases.

A key amendment of the IHR (2005) requires States Parties to establish a National IHR Authority (NIA), tasked with coordinating the implementation of the regulations.^2^ Strong multisectoral coordination is needed to effectively implement core capacities at the human–animal–environment interface; to have the financial systems to reliably fund prevention, preparedness, response and recovery activities; to manage and reduce the risk of chemical, radiation and food safety incidents; and to maintain whole-of-government and whole-of-society coordination and policies for efficient response to public health emergencies.^3^ This highlights that strengthening core capacities requires not only technical enhancements but also strong political commitment and effective collaboration across multiple sectors.

Importantly, the role of the NIA is different to that of the National IHR Focal Point (NFP). NIAs will be mandated to drive policy, resource allocation and multisectoral engagement, while NFPs focus primarily on communication between WHO and States Parties. A fully operational NFP ensures seamless communication with international partners and guarantees that health security information—including notifications, verifications and reports—is conveyed accurately and promptly.^4^ This precise and timely exchange of information is crucial for the early detection of and effective response to public health risks and emergencies, and is in itself a core capacity.^5^

Based on self-reporting, the State Party Self-Assessment Annual Reporting (SPAR) tool is used to systematically report progress on the level of IHR core capacity implementation. For 15 core capacities, States Parties report where they have limited or no capacity (Level 1) and where they have advanced or sustained capacity (Level 5).^6^ The core capacities span critical domains, such as surveillance, laboratory, risk communication and community engagement, and financing. States Parties assign performance scores—typically ranging from 0 to 100%—using a standardized methodology, with each indicator assessing specific operational aspects. Monitoring SPAR results helps identify gaps and prioritize capacity-building efforts.^7^

As States Parties are expected to establish and maintain core capacities required under the IHR, there is a need to understand the current status of those capacities. This can inform the context and priorities for the establishment or continued existence of an NIA, as well as priority actions for future robust implementation of the core capacities. This paper provides an analysis of the core capacities in the Western Pacific Region. The findings will help States Parties to consider future capacity-strengthening priorities and the priorities for IHR amendments implementation including the role and entity designated as the NIA.

## METHODS

Each year, States Parties are required to report the status of IHR core capacities using SPAR, a standard tool introduced in 2021.^8^ For this analysis, 2021–2023 scores of States Parties in the Western Pacific Region were obtained from the eSPAR platform, which is publicly available online. To describe the status of IHR core capacities, the average SPAR scores for each of the 15 indicators were calculated, rounded and colour-coded for the years 2021, 2022 and 2023. The colour coding represents the status of core capacity implementation, where higher scores indicate greater capacity based on self-reporting. The colour scheme is as follows: red (0–20), orange (21–40), yellow (41–60), light green (61–80) and dark green (81–100).

For the analysis, States Parties were categorized into three groups—high-income, low- and middle-income, and Pacific island—based on geographic and economic characteristics, referencing 2023 World Bank classifications.^9^ A radar chart was utilized to visualize each overall core capacity score (abbreviated as C1–C15). States Parties with missing data were excluded from analysis.

## RESULTS

More States Parties reported on their IHR core capacities in 2023 compared to earlier years (Table 1). All States Parties reported at least once during the 3-year period, with

**Table 1.**
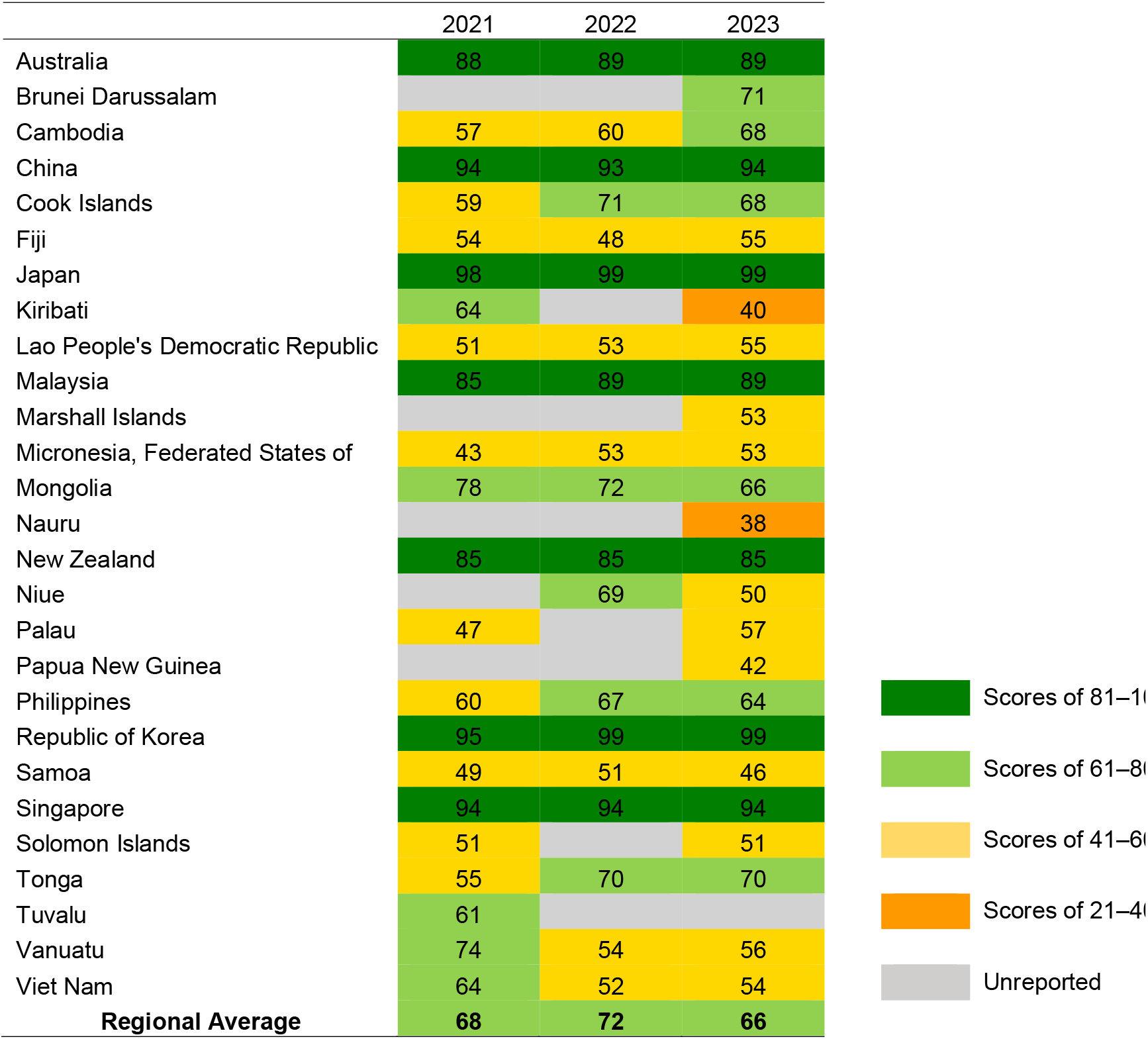
Average score for IHR core capacities among States Parties in the Western Pacific Region, 2021–2023 (%)

19 reporting every year. The regional average score increased from 68% in 2021 to 72% in 2022, then declined to 66% in 2023 (Table 1).

Seven States Parties (Australia, China, Japan, Malaysia, New Zealand, the Republic of Korea and Singapore) maintained strong and stable scores, consistently exceeding 85%. Nine States Parties (Cambodia, Cook Islands, Kiribati, the Federated States of Micronesia, Niue, Palau, Tonga, Vanuatu and Viet Nam) exhibited large fluctuations (10 points or more) in yearly scores. One State Party (Mongolia) reported a slight decline, and two (Cambodia and Lao People’s Democratic Republic) reported steady increases in core capacities across the years.

Among the 26 of 27 States Parties that reported in 2023 (Fig. 1), good capacity (≥60% score) was reported for laboratory (C4), surveillance (C5), health emergency management (C7), health service provision (C8) and risk communication and community engagement (C10). The biggest core capacity gaps were reported for zoonotic diseases (C12), food safety (C13), chemical events (C14) and radiation emergencies (C15). Scores varied across income and geographic groupings, where high-income States Parties generally posted higher overall scores, Pacific island States Parties tended to report lower capacities, and low- and middle-income States Parties reported gaps in financing (C3) and infection prevention and control (C9).

**Fig. 1.**
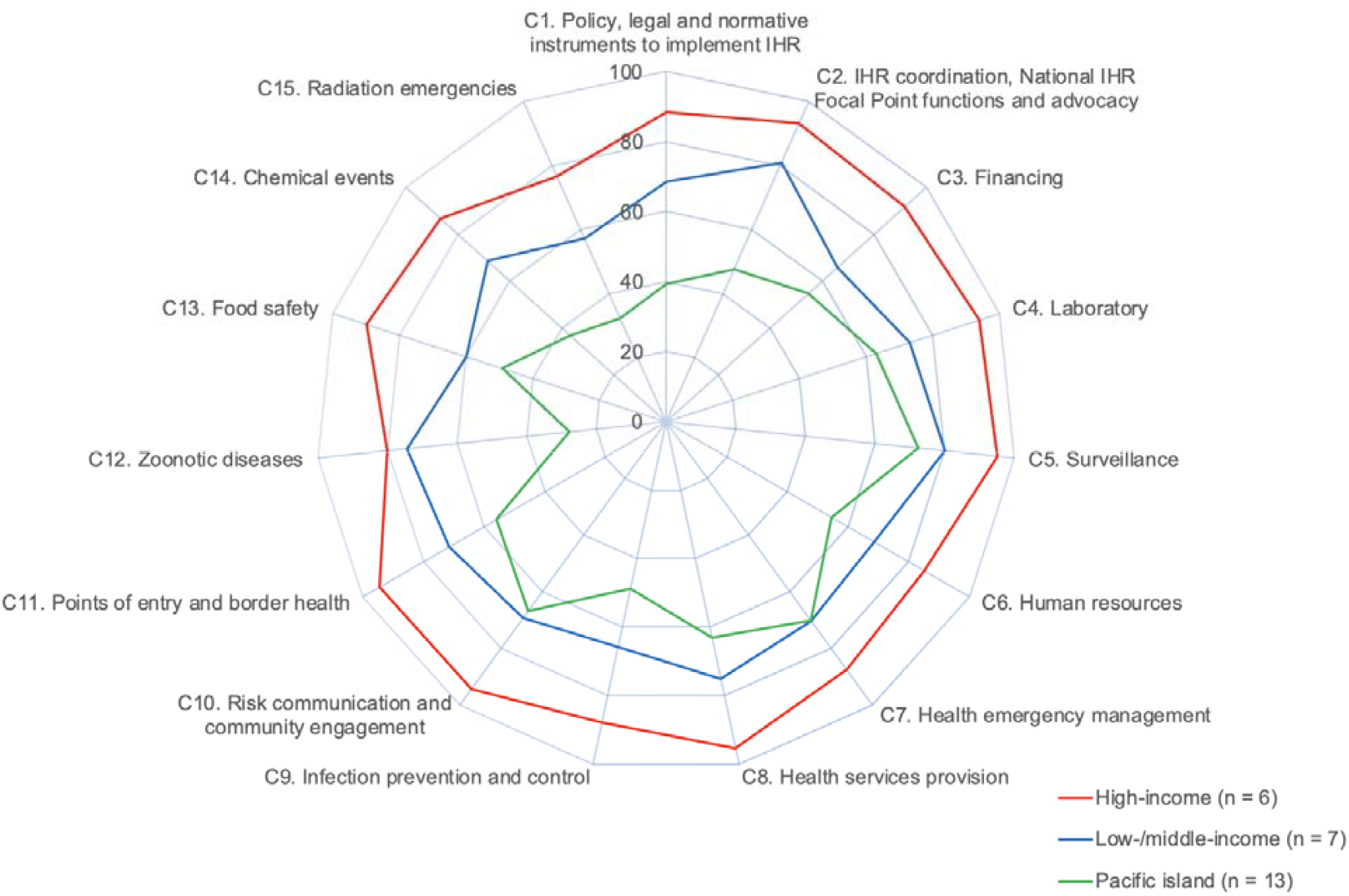
Average IHR core capacity scores in the Western Pacific Region, 2023 States Parties in the Western Pacific Region are categorized into three groups based on geographic and economic characteristics, referencing 2023 World Bank classifications:^9^ High-income: Australia, Brunei Darussalam, Japan, New Zealand, Republic of Korea, Singapore. Low-/middle-income: Cambodia, China, Lao People’s Democratic Republic, Malaysia, Mongolia, Philippines, Viet Nam. Pacific islands: Cook Islands, Fiji, Kiribati, Marshall Islands, Federated States of Micronesia, Nauru, Niue, Palau, Papua New Guinea, Samoa, Solomon Islands, Tonga, Vanuatu. Tuvalu did not report data for 2023 and was excluded from analysis.

## DISCUSSION

The status of IHR (2005) core capacities among States Parties in the Western Pacific Region reflects the diversity in national systems, resources and contexts. While some have made notable progress in areas such as surveillance, laboratory services and emergency management, all States Parties have opportunities to further strengthen specific domains—particularly food safety, zoonotic disease control and sustainable financing. Sustained investment and coordination are critical for ensuring that all States Parties can effectively prevent, detect and respond to public health threats.

Multiple Joint External Evaluations (JEEs) conducted in the region provide additional insights into areas requiring attention. For instance, in the Pacific islands, States Parties have highlighted persistent workforce shortages, limited surge capacity, and the critical need for multisectoral coordination mechanisms to rapidly mobilize external support in response to chemical/radiation emergency situations or other acute public health hazards. In particular, health workforce shortages and geographic dispersion continue to pose significant challenges for Pacific island States Parties.^10–15^

Significant progress has been made by States Parties in strengthening core capacities. For example, by establishing and reinforcing emergency medical teams (EMTs), States Parties have bolstered their shared ability to rapidly respond to disease outbreaks and disasters—thus reinforcing IHR core capacities critical for effective health emergency management. Since the inception of the EMT Initiative in 2010 following the devastating Haiti earthquake, 16 of the 51 WHO-classified EMTs (31%) established are in the Western Pacific Region. Alongside these EMTs classified for international response, nearly every Member State in the Region has established a national EMTs or is in the process of doing so. This means that nearly all States Parties have domestic EMTs ready to surge or can offer assistance to others in times of crisis. In recent years, EMTs from the Western Pacific have deployed to provide rapid clinical care in disasters, outbreaks and mass gathering events, but they have also helped build local capacities through joint training and full-scale simulation exercises. Their presence and coordinated action facilitate knowledge transfer and enhance emergency management capacities. These measures acknowledge that while many States Parties have relatively small health systems, they can leverage regional solidarity and external technical assistance to address chemical, biological and radiological incidents more effectively (Casey ST, Noste E, Cook AT, Muscatello D, Heslop DJ. Emergency medical teams in WHO’s Western Pacific Region. 2025 [Submitted to Western Pac Surveill Response J]).^16–20^

A similar story can be told about the regional uptake of the Global Outbreak Alert and Response Network (GOARN), where 80 of the 320 (25%) participating institutions globally are from the Western Pacific Region. Nearly 90 GOARN missions were conducted in the Region during the COVID-19 pandemic, and the mechanism was more recently used to respond to measles events to bolster clinical management and infection prevention and control. The experts deployed not only supported immediate needs but provided training to prepare health systems for future outbreaks. It is worth noting that the investments for EMT, GOARN and other surge mechanisms come from within the Region itself, where donors and countries act in solidarity.^21^ These mechanisms not only strengthen regional response capacities but also enable cross-border collaboration, ensuring that expertise and resources can be mobilized swiftly within and beyond the Western Pacific Region when needed.

Zoonotic disease control stands out as another key example of capacity building in the Region. Fourteen out of 27 States Parties in the Western Pacific Region have established multisectoral coordination mechanisms integrating the human health, animal health and environmental health sectors to detect and contain zoonotic threats more efficiently.^22,23^

Viet Nam’s integrated response to a recent outbreak of foodborne illness in 2024 exemplifies the tangible impact of such multisectoral planning: laboratory capacities, epidemiological surveillance and risk communication measures were activated in concert, preventing further spread and enabling prompt public advisories.^24^ The International Food Safety Authorities Network (INFOSAN) further accelerates information sharing and collective action, ensuring that food safety incidents can be addressed swiftly and collaboratively. As of December 2024, 27 States Parties (100%) in the Western Pacific Region have INFOSAN contact points. By engaging into this network, States Parties can bridge capacity gaps, share critical data and coordinate timely responses to protect public health.^25^ Investment in food safety has major collateral benefits for strengthening antimicrobial resistance monitoring, genomic surveillance and food supply chains. Future regional efforts in food safety incident management, especially maintaining sustainable funding for said efforts, will be critical.

Despite notable progress in these areas, many States Parties still face challenges in managing hazards—especially chemical, radiological and food safety events—that require robust multisectoral coordination. The recent amendments to the IHR (2005) underscore the importance of strengthening core capacities to address evolving public health threats, going beyond the maintenance of existing standards to require rapid detection, assessment, reporting and response. Establishing an NIA provides an opportunity to further strengthen multisectoral collaboration, resource integration and international collaboration. An effectively empowered NIA can coordinate these efforts by engaging multiple stakeholders and driving both whole-of-government and whole-of-society approaches. A well-resourced NIA can optimize resource allocation, streamline decision-making and foster transparent information sharing, thereby making steady progress toward more robust IHR implementation. In turn, this helps address persistent gaps in areas such as food safety, chemical and radiological preparedness and risk communication.

Consequently, regional coordination becomes imperative. Establishing a regional health emergency forum or council could foster political commitment and joint action, strengthening collective resilience. Where establishing complete domestic capacity is not practical—especially for certain chemical or radiation events—States Parties may benefit from existing regional networks and technical support arrangements, which allow resource-limited States Parties to leverage international expertise as needed.

These considerations are especially relevant given the low incidence but potentially high impact of certain incidents, such as chemical or radiation events in many island settings, as well as the prohibitive costs of maintaining full-scale national response systems. As a result, several Pacific island States Parties rely on formal agreements with larger neighbours or regional hubs for technical expertise and laboratory analyses. Such arrangements underscore the need for well-defined protocols and multisectoral mechanisms to rapidly mobilize external support in the face of chemical spills, radiation leaks or other complex hazards.

Overall, strengthening data collection and information-sharing practices is crucial for gaining a comprehensive understanding of progress in core capacities. It is important to note that the IHR core capacity data reported to WHO each year are based on self-reports, which may under- or overestimate capacities. Nevertheless, the reporting is an important step in shared knowledge and transparency that can guide investments and strategic implementation. Incorporating qualitative assessments such as JEEs alongside SPAR reporting can highlight nuanced challenges and opportunities. Regular SPAR reporting and proactive exchange of experiences among States Parties promote transparency, identify best practices and facilitate collective improvement. The NIA will play a critical role in furthering the implementation of the IHR core capacities in States Parties, and the opportunity for international coordination is highlighted in this analysis whereby States Parties can leverage each other’s capacities in solidarity. The Region’s diverse capabilities and historically interwoven public health implementation pave the way forward for stronger health security.

## Data Availability

This regional analysis consists of a review and synthesis of publicly available public health data. It does not involve human participants, identifiable personal data or interventions. Based on organizational ethical review policies, such activities do not require formal ethics approval.

